# Incorporating Nanopore Sequencing into a Diverse Diagnostic Toolkit for Incontinentia Pigmenti

**DOI:** 10.1101/2023.09.26.23295778

**Authors:** Simone Ahting, Denny Popp, Henry Oppermann, Vincent Strehlow, Maria Fasshauer, Bernt Popp, Maike Karnstedt, Isabell Schumann

## Abstract

**Background:** Incontinentia pigmenti (IP) is a rare, hereditary multisystemic disorder affecting 1.2 in 100,000 live births, predominantly females. Conventional genetic analyses through short-read sequencing are complicated in case of IP due to the presence of a highly homologous pseudogene. Traditionally, long-range PCR is employed in order to overcome this challenge, however, detection of skewed X-Inactivation can also aid in correctly assign a variant to *IKBKG*.

**Methods:** We employed a comprehensive multi-method approach, incorporating whole-exome sequencing (WES), long-range PCR, RT-PCR, X-inactivation analysis, and nanopore sequencing of genomic DNA, to identify and accurately phase a small heterozygous deletion NM_001099857.5:c.363_367del, p.(Leu122Glyfs*14) to the *IKBKG* gene in a family affected with IP.

**Results:** The deletion was initially detected through WES and skewed X-inactivation was observed in both the proband and her mother. Long-range PCR specific to *IKBKG* was utilized to verify that the variant is located in *IKBKG* and not in its highly homologous pseudogene. On RNA level, the variant was undetectable, suggesting nonsense-mediated decay (NMD) of the transcript containing the variant. We further utilized nanopore sequencing not only to pinpoint and accurately map the variant to the *IKBKG* gene but also to analyze methylation status of both alleles. This allowed us to confirm the skewed X-inactivation, with the variant-carrying allele found to be predominantly inactivated.

**Conclusion:** Nanopore sequencing serves as a valuable tool in genetic diagnosis, enabling the precise localization of the variant in either the gene or the pseudogene. Furthermore, in females with skewed X-inactivation, this method facilitates the determination of whether the variant is predominantly located on the activated or inactivated X-chromosome.

## 1. Introduction

Incontinentia pigmenti (IP; Bloch-Sulzberger-syndrome, OMIM #308300) is a rare hereditary multisystemic neuroectodermal disorder caused by pathogenic variants in the *IKBKG* gene (formerly known as *NEMO* (nuclear factor-kappa B (NF-kB) essential modulator)) on chromosome X. It occurs in 1.2 out of 100,000 live births^1^, mainly affects females and is generally lethal in males during embryogenesis^2^, with the rare exception of cases with somatic mosaicism or XXY karyotype^3^. IP is characterized by typical skin manifestations along the Blaschko lines shortly after birth that can be divided into four successive inflammatory stages from blisters and pustules on the extremities (stage 1), to verrucous lesions (stage 2), hyperpigmentation (stage 3) and hypopigmentation (stage 4) that often persists into adulthood^4^. Ophthalmologic, odontologic and neurologic impairment can occur additionally as systemic manifestation of IP. In females with heterozygous pathogenic *IKBKG* variants, cells expressing the mutated allele are usually selected against early in life, leading to extremely skewed X-chromosome-inactivation^5^ (XCI) that can serve as a diagnostic criterion for IP. This elimination of mutant allele-expressing cells is due to the vital role of *IKBKG* in the activation of NF-kB pathways that are crucial for many physiological functions, such as immune, inflammatory, anti-apoptotic and developmental pathways^6^. Cells lacking the *IKBKG*-encoded NEMO/IKKγ protein due to loss-of-function variants show increased sensitivity to pro-apoptotic stimuli ^7^ hence driving skewing of XCI. Traditionally, this can be investigated through analysis of methylation status on CpG sites of two polymorphic X-chromosomal repetitive elements near the *AR* and the *RP2* locus, that can be used for separation of the parental alleles^8,9^.

The *IKBKG* gene (NM_001099857.5) on the Xq28 locus is 23 kilobases (kb) long and consists of nine coding-exons (exons 2-10), an additional four alternative non-coding first exons (1A-D), as well as two promoters. The uni-directional promoter A directs transcription of exons 1A and 1D and is located in intron 2 of the *G6PD* gene that is situated on the opposite strand. Promoter B is a strong bidirectional promoter driving transcription of exon 1B and 1C as well as *G6PD* itself^10^. A nonfunctional second copy of *IKBKG* with >99% sequence identity^11^ comprising the exons 3-10 makes up the non-processed, non-transcribed *IKBKG* pseudogene *IKBKGP1*. Together, they are part of a 35.7 kb segmental duplication that consists of two low copy repeats (LCR1/2) adjacent and in opposite orientation, one covering *IKBKG*, the other one *IKBKGP1*^10^ (Fig.1). The most common pathogenic variant accounting for 70-80% of IP cases is a large deletion spanning the exons 4-10, termed *IKBKG*del^12^. Other variants pathogenic for IP have been reported and include small delins (54%) and single-nucleotide substitutions (46%). The latter are mostly variants that lead to premature termination codons (PTCs), either through nonsense or frameshift changes, while only 15% are missense variants^12^. In fact, owed to the high homology of the *IKBKG* and the *IKBKGP1* loci, *intra*-locus genomic rearrangements are frequent and have been shown to render the IP locus susceptible to novel pathological alterations^10^. Due to the high similarity of the *IKBKG* and the *IKBKGP1* locus, detection of *IKBKG* variants using short-read sequencing is hindered, since the presence of *IKBKGP1* affects read-depth, mapping quality and alignment, increasing the risk for false-positive or false-negative results^13^. Therefore, so far, Sanger-Sequencing using long-range PCRs has been used as the method of choice in IP diagnostics^14^.

**Figure 1:**
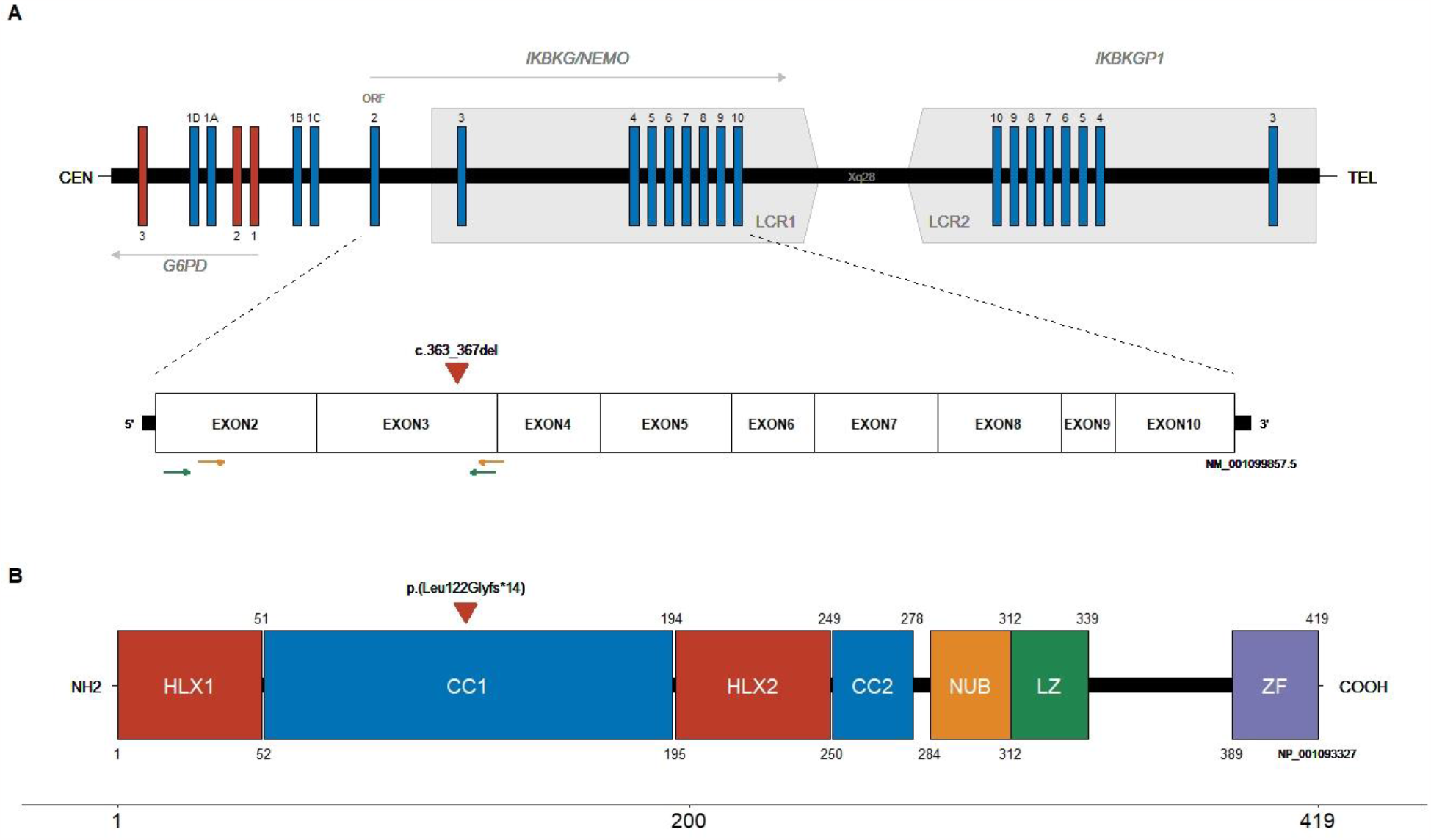
The *IKBKG* locus on chromosome X (adapted from Pescatore *et al*., 2022 ^36^). (**A**) Schematic representation of the Xq28 locus including *IKBKG*, its pseudogene *IKBKGP1* and the NM_001099857.5 transcript. The square grey arrow boxes represent the Low Copy Repeats (LCR1/2) of the segmental duplication, the exons of *IKBKG/IKBKGP1* are depicted in blue, the exons of *G6PD* in red. The *IKBKG* gene incorporates nine coding exons and four alternative non-coding exons (1A-1D) that overlap with the locus of *G6PD*. Translation initiation site is located at the beginning of exon 2, marked here with ORF. The transcript is composed of the exons 2-10, the identified variant c.363_367del in exon 3 of *IKBKG* is marked with a red triangle. RNA-primerpairs used for confirmation of variant location are shown as green and orange arrows (Primerpair B = orange arrows, Primerpair C = green arrows, see Suppl. Table 2). (**B**) Schematic representation of the IKBKG protein, its domains as well as indication of the identified variant p.(Leu122Glyfs*14) in NP_001093327 (419 aa). Domains are as follows: HLX1 – *Helical domain*; CC1 - *Coiled Coil*; HLX2 - *Helical domain*; CC2 - *Coiled Coil*; NUB - *NEMO Ubiquitin Binding*; LZ - *Leucin Zipper*; ZF – *Zinc Finger;* aa ranges are indicated below and above the domain.

In our present study, we describe a female individual with characteristic phenotype of IP and a similarly affected mother. We applied whole-exome-sequencing (WES), Sanger-Sequencing, X-inactivation analysis, RT-PCR and long-read amplicon sequencing to confirm a novel frameshift variant in *IKBKG* in the affected family. Furthermore, we utilized Oxford Nanopore Technologies (ONT) sequencing not only to differentiate whether the variant is located in the gene or pseudogene but also succeeded in ascertaining whether the variant was located on the predominantly active or inactive X-chromosome by analyzing the methylation frequency at the two *IKBKG* promoters.

## 2. Material & Methods

### 2.1. Ethics and consent

This study adheres to the principles set out in the Declaration of Helsinki. The study was approved by the Ethical Committee of the Medical Faculty of the Leipzig University and the authors received and archived written consent of the affected individuals and/or their legal guardians to publish genetic and clinical data as well as photographs.

### 2.2. Exome sequencing

Genomic DNA was isolated from EDTA-blood samples of the index and parents using the MagCore HF16 Plus Nucleic Acid Extractor, DNA concentration was measured using a NanoDrop 2000 (Thermo Scientific). Enrichment and library preparation for exome sequencing was performed using the TWIST Human Core Exome Kit (TWIST Bioscience, San Francisco, CA, USA). Libraries were sequenced with 150 basepairs (bp) paired-end reads on a NovaSeq 6000 system (Illumina, Inc., San Diego, CA, USA). On average coverage of targeted genomic regions was >100x with 97% covered at least 10x. Raw data was processed using varfeed followed by a tertiary analysis with the browser-based genomics software Varvis (Limbus Medical Technologies GmbH, Rostock, Germany). The variant in *IKBKG* (NM_001099857.5, *MANE*-select) was classified according to the ACMG criteria and latest recommendations^15^, the ACGS Best Practice Guidelines 2019^16^ and the ClinGen Sequence Variant Interpretation Recommendations for PM2 - Version 1.0^17^.

### 2.3. Sanger sequencing

Bidirectional Sanger sequencing for confirmation and segregation analysis of the parents was performed on the Applied Biosystems 3500 Genetic Analyzer (Thermo Fisher Scientific Inc., Waltham, Massachusetts, U.S.), using two sets of primers binding to regions in exon 3 or adjacent introns (Suppl. Tab. 1). The identified variant was submitted to ClinVar^18^. Bioinformatic analysis was done using the Sequence Pilot Software (JSI medical systems, Ettenheim, Germany).

### 2.4. RNA analyses

For identification of the variant in *IKBKG* on RNA level, RNA was isolated from peripheral blood lymphocytes using the PAXgene Blood System (Becton Dickinson, Franklin Lakes, NJ, U.S.) of the individual and mother. Complementary DNA (cDNA) was generated using PrimeScriptTM RT Master Mix (TaKaRa Bio Europe SAS, Saint-Germain-en-Laye, France) according to manufacturer’s instructions. RT-PCR was conducted followed by bidirectional Sanger sequencing of parts of the exons 2-4 (Suppl. Tab. 2) on the Applied Biosystems 3500 Genetic Analyzer (Thermo Fisher Scientific Inc., Waltham, Massachusetts, U.S.) and PCR product was visualized by gel electrophoresis. Bioinformatic analysis was done using the Sequence Pilot Software (JSI medical systems, Ettenheim, Germany).

### 2.5. Validation of variant by long-range-PCR

For validation of the variant as well as differentiation between *IKBKG* and *IKBKGP1*, long-range PCR of the *IKBKG* locus in the index patient, amplification of the respective exon as well as sequencing was performed by congenics Humangenetik MVZ in Osnabrück, Germany.

### 2.6. X-Inactivation

X-inactivation testing on EDTA whole blood was recommended after variant identification for both the index and the mother. To this end, XCI status was evaluated through digest by methylation-sensitive restriction enzyme *Hpa*II followed by PCR-amplification of digested and undigested genomic DNA using primers for the highly polymorphic CAG-repeat locus in the *AR*-gene^9^ as well as the extragenic tandem-repeat-GAAA-locus 5’ of the RP2 promoter^8^. Products were separated using an Automatic Genetic Analyser and methylation status was calculated as previously described^19^. Skewing is present when the predominant allele constitutes more than 74% of all alleles, a percentage between 90% and 100% is considered extreme skewing.

### 2.7. Nanopore sequencing

DNA (3 μg) from EDTA-whole-blood was prepared for ONT sequencing using the Ligation Sequencing Kit (SQK-LSK114, Oxford Nanopore Technologies) according to the manufacturer’s instructions with the following adjustments: Incubation time of the end-prep reaction was increased to 15 min at 20 °C and 15 min at 65 °C. Incubation time of adapter ligation reaction was adjusted to 30 min at room temperature. The library (300 ng) was loaded onto a PromethION flow cell type R10.4.1 and sequenced on a P2Solo device (ONT). The sequencing run was performed using adaptive sampling targeting the q-arm of the X chromosome as well as the *RP2* locus on the p-arm with 50 kb flanking region up- and downstream. The flow cell was washed twice, once after 18 h and again after a further 29 h of sequencing using Wash Kit EXP-WSH004 (ONT) according to the manufacturer’s instructions. After each washing step, another 300 ng of DNA was loaded onto the flow cell.

Raw sequencing data (fast5 files) was basecalled and mapped against the reference human genome hg38 by guppy v6.5.7 (ONT) using a methylation-aware basecalling model with enabled read splitting, adapter trimming and calibration strand removal. In total, 38.7 Gb of sequencing data was obtained with an on-target read length N50 value of 19 kb. After exclusion of alignments with a mapping quality below 20, variants were called using clair3 v1.0.4^20^ and phased using whatshap v1.7. Whatshap was further used to assign tags to the reads based on haplotypes and to partition the alignment file into separate haplotypes. For each haplotype, the methylated fraction of the promotor region of the *IKBKG* gene (chrX:154546678-154547921) was calculated and visualized using methylartist v1.2.7^21^.

## 3. Results

### 3.1. Clinical description

The female individual was first brought in for evaluation very early in her life. Pregnancy was unremarkable and she was delivered spontaneously. Birth weight, birth length and head circumference were all within the normal range. Initially she showed yellow purulent skin lesions at her left wrist and forearm as well as gluteal efflorescences. Due to breathing difficulties and cyanosis she was admitted to neonatal intensive care unit and was discharged from the hospital without breathing difficulties shortly after. One day later she was admitted again due to poor drinking, hypothermia and circulatory centralisation suspected to have a neonatal infection. Physical examination showed no signs of infection, but in the course, her skin showed yellow-white incrusted blisters, papules and erythema on her upper and lower extremities. A consulted dermatologist discussed the possibility of IP. The skin lesions healed a few months later.

Speech and motor development were normal at the time of her last examination in our genetic counselling in early infancy. At her physical examination stages, one and two of her skin manifestation had already been regressed and she showed very faint white maculae along the Blaschko lines in groin and knee pit. She had no other systemic involvements at that time. Both the individual’s mother as well as maternal grandmother also reported to have had skin lesions shortly after birth, but had no other symptoms throughout their lives. Only the index and the mother were available for testing (see Suppl.Fig. S1 & Fig. 2 (available on request)).

**Figure 2:** Individuals’ skin manifestations: Pictures regarding the individuals’ phenotype are available on request with the corresponding author.

For a detailed description of the individuals’ phenotype and medical history, please contact the corresponding author.

### 3.2. Molecular analysis

We performed WES to analyze comprehensively both single nucleotide variants (SNVs) as well as larger copy number variations (CNVs) and simultaneously evaluate differential diagnoses. We found a heterozygous likely pathogenic 5 bp deletion GRCh38/hg38: chrX:g.154556340_154556344del, NM_001099857.5(*IKBKG*):c.363_367del, p.(Leu122Glyfs*14) in exon 3 of the *IKBKG* gene (Fig. 1). Segregation using Sanger sequencing confirmed the finding and revealed the variant to be maternally inherited. Due to methodical limitations of short read sequencing, we were not able to determine whether the variant was actually located on the *IKBKG* gene or its highly homologous pseudogene *IKBKGP1* and could therefore not conclusively confirm suspicion of IP (Suppl. Fig. S2). Subsequently performed RNA analysis was meant to address this since *IKBKGP1* is not transcribed. Hence, only *IKBKG* transcripts would be amplified. However, neither of the two employed primer pairs for RNA analysis (Suppl. Tab. 2) amplified sufficient quantities of the variant-carrying transcript for detection. Consequently, sequencing revealed only PCR products from the wild-type *IKBKG* transcript.

Next, we checked for skewed XCI in the blood of the proband and her mother. Pathogenic variants in *IKBKG* usually lead to skewed XCI due to negative selection against cells expressing the mutated allele^5^. XCI analysis investigating polymorphic repetitive elements in *AR* and *RP2* strengthened the suspicion of IP by showing extreme skewing in the mother of the proband with a ratio 95:5 and a skewed XCI in the proband with a ratio of 85:15, consistent with the diagnosis. Subsequently conducted long-range PCR followed by sequencing of the probands DNA confirmed the location of the variant on the *IKBKG* gene thereby affirming the diagnosis of IP in the probands family.

We also incorporated ONT sequencing as part of the investigation process. To this end, we applied adaptive sampling targeting the q-arm of chromosome X in the proband, thereby verifying the variant location in the *IKBKG* gene (Fig. 3). Furthermore, we used the ONT-generated 5mc methylation information to infer whether the variant in *IKBKG* is preferentially located on the activated or inactivated chromosome X through analysis of the haplotype-specific methylation of the two *IKBKG* promoters (see Fig. 4). The methylated fraction in the promoter region of haplotype 1 (paternal), which is designated as wild-type since it does not carry the variant in *IKBKG*, was 7.9%. In contrast, the methylated fraction of haplotype 2 (maternal) carrying the variant in *IKBKG*, was 71.7%, suggesting a higher likelihood of being present on the inactivated X chromosome. These findings validate the previously mentioned skewed X-inactivation pattern in the individual and further confirm the mutated allele as the one preferentially undergoing inactivation. A methylation analysis of the *AR* and the *RP2* loci also confirmed the skewed XCI (Suppl. Fig. S3).

**Figure 3:**
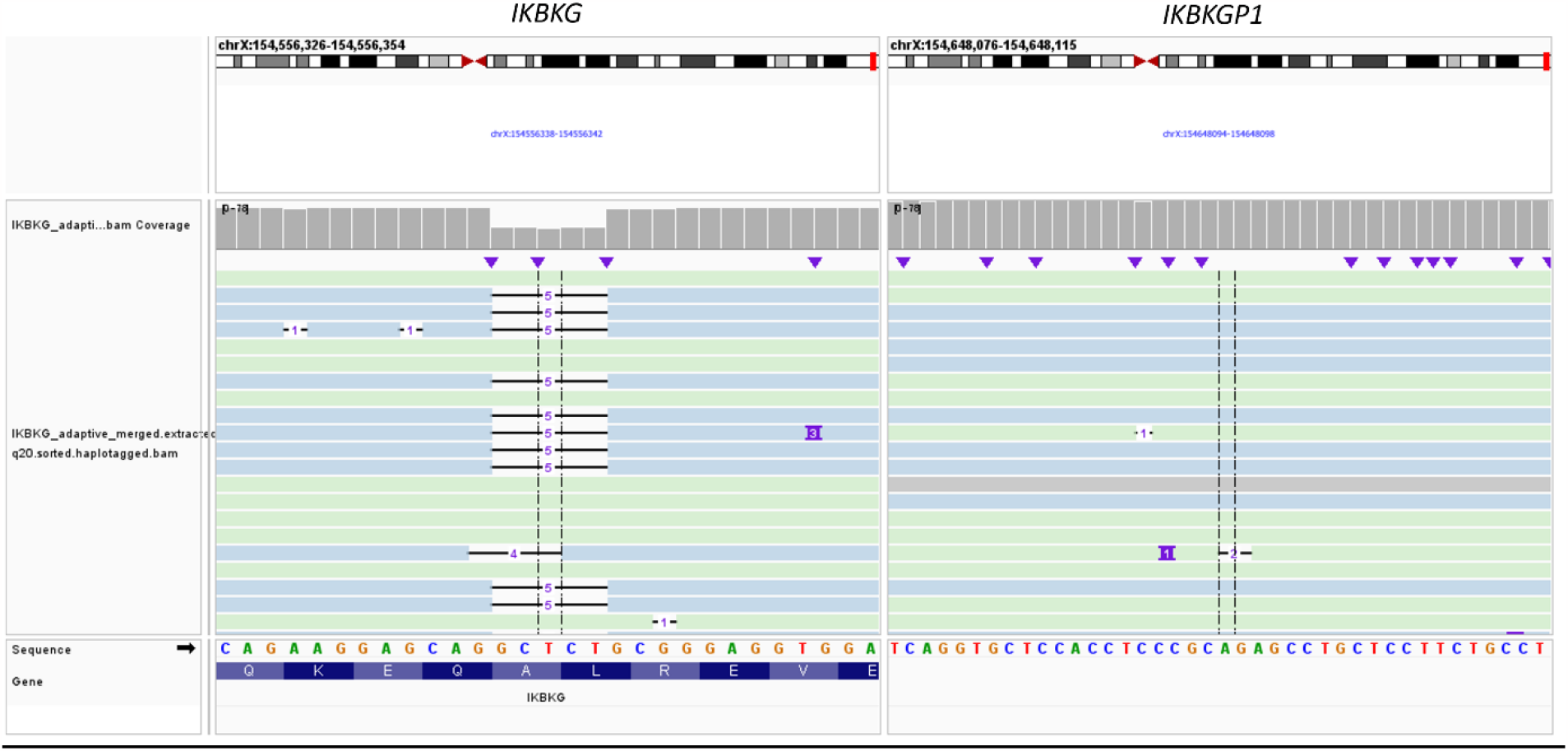
IGV screenshot of aligned Nanopore reads: The genomic region around the variant c.363_367del, p.(Leu122Glyfs*14) in the *IKBKG* gene (left panel) as well as the homologous region in the pseudogene *IKBKGP1* (right panel) in which none of the reads show the variant of interest. Reads are colored according to haplotypes.

**Figure 4:**
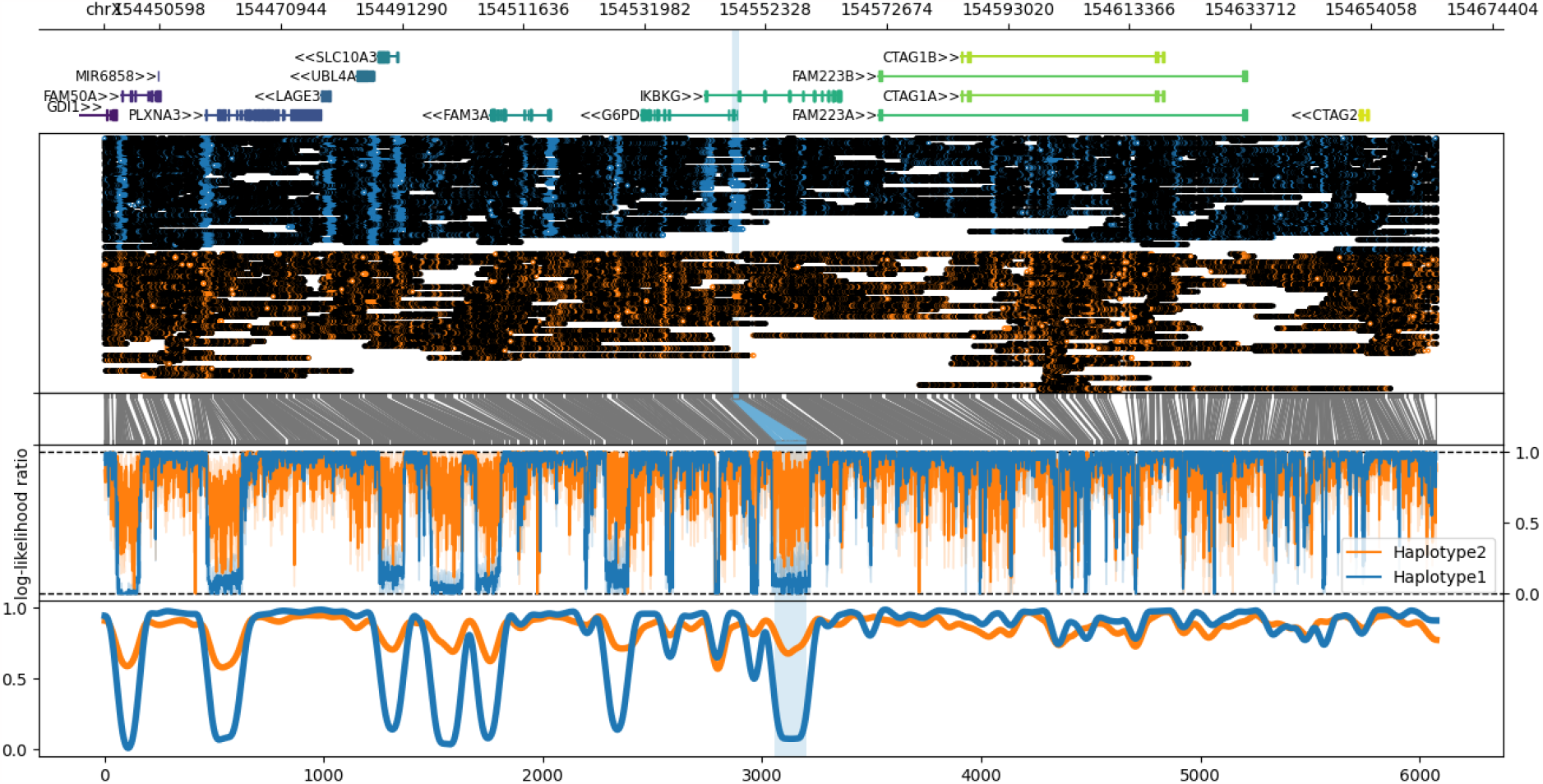
Haplotype-specific methylation profiles of the *IKBKG* locus showing increased methylation of the variant-carrying maternal allele (haplotype 2, orange) compared to the wild-type paternal allele (haplotype 1, blue). From top to bottom this plot shows a gene track, haplotype-specific methylation calls (black color coded) relative to aligned read positions, a translation from genome space into a modified base space consisting only of instances of the methylated motif, the haplotype-specific methylation statistic (log-likelihood ratio: 1: likely methylated, 0: likely unmethylated) and a smoothed sliding-window plot showing the methylated fraction across the region (*IKBKG* gene ± 100 kb). The promoter region of the *IKBKG* gene is highlighted in light blue.

## 4. Discussion

The hallmarks of IP encompass the cutaneous manifestations generally characterized by four distinct stages that can overlap or may not develop at all in some patients^24^, while other organs may be affected later in life. Here we describe the case of a young female individual carrying a novel maternally inherited small deletion in *IKBKG*. Both the individual and the mother showed no signs of hyperpigmentation along the Blaschko lines and hypopigmented areas were very faint. The former is surprising, since hyperpigmented areas of the skin develop in as much as 98% of affected females and can persist up to the fourth decade of life^25^, while hypopigmented skin areas have been suggested to be dramatically underreported in the past^24^. Furthermore, studies on genotype-phenotype correlations are very rare and suggest that variant type, affected domain, status of XCI and genomic background may all contribute to the high phenotypic variability reported in IP patients ranging from mild skin alterations to severe central nervous system (CNS) abnormalities^7^. Interestingly, loss-of-function variants have been shown to coincide a higher likelihood of skewed or extremely skewed XCI^26^ and skewing seems to negatively correlate with disease severity and CNS involvement^27^. Also, hypomorphic variants (variants that preserve some protein activity) show a greater involvement of different tissues compared to more severe protein function abolishing variants^26^. These findings support the effect of the small deletion we report here and offers an explanation to the rather mild, single-organ phenotype of the family. In confirmation, XCI status of the family showed extremely skewed XCI in the mother (95:5) and skewed XCI in the index patient (85:15), consistent with the notion that skewing is known to increase with age^28^ and consistent with the above-mentioned correlations.

Diagnostic strategies for IP still heavily rely on long-range PCR and Sanger sequencing as the gold standard method for testing^14^. However, broad screening technologies such as WES are exceedingly employed in routine genetic laboratories. This short-read based capture probe data analysis makes reliable discrimination between highly similar genes and pseudogenes difficult and identified variants need to be validated using an independent method. Recent innovations have tried to bypass this by optimizing the bioinformatics pipeline in order to mask the *IKBKGP1* pseudogene allowing only for detection of variants in *IKBKG*^29^. This approach could potentially serve as an alternative to the traditional long-read Sanger sequencing. In addition, since IP is an X-linked disorder, analysis of skewed XCI can serve as a diagnostic criterion, aiding in the determination of whether or not a variant might be causative for the patients’ symptoms. Traditional analysis of fragment lengths after methylation sensitive restriction enzyme digest at the *AR* and *RP2* loci is frequently employed for analysis of skewed XCI. Most recently, this has also been achieved using ONT sequencing after CRISPR-Cas9 enrichment, demonstrating a series of advantages towards traditional diagnostic strategies and reliably and reproducibly determining the XCI status in multiple females^30^. Since ONT sequencing relies on long-reads and allows for pinpointing a variant’s location within a gene, even in the presence of closely related pseudogenes, it can facilitate both variant mapping and XCI analysis, even extending to the precise assignment of the variant to the predominantly active or inactive X-chromosome. This combination of features might render ONT sequencing a potent tool, particularly valuable in cases of X-linked disorders characterized by recognizable XCI skewing.

Here, WES revealed the presence of a small deletion in either *IKBKG* or *IKBKGP1*. Since mapping was inconclusive, and IP was specifically suspected by the physician, we visually explored the *IKBKG* locus using the Integrative Genomics Viewer (IGV). Still, we were unable to assign the variant to either *IKBKG* or *IKBKGP1*. RNA-analysis was meant to address this issue, since *IKBKGP1* is not transcribed, however, we were not able to amplify the variant carrying transcript and sequenced wild-type transcripts only. At the time, we were not certain if this was due to the variant being located on the *IKBKGP1* locus or due to nonsense-mediated mRNA decay (NMD; a physiological surveillance pathway degrading mutant mRNAs harboring PTCs^31,32^) of *IKBKG* transcripts carrying the variant. Due to the inflicted frameshift and ultimate PTC of the detected variant as well as its location within the transcript, rapid degradation by NMD is highly likely, hence preventing it from detection via mRNA-analysis through Sanger sequencing. Strategies to circumvent this diagnostic gap have been reported previously, mainly relying on pharmacological inhibition of NMD^33^ that can be adapted to the requirements of diagnostic laboratories^34^. Given that XCI is skewed in the proband at an 85:15 ratio, transcription of the transcript harboring the variant is anticipated to occur in only 15% of cells, resulting in an allelic fraction (AF) of approximately 7.5%. Taking into account the effect of NMD, detection of the variant via Sanger sequencing becomes highly improbable, considering its established detection threshold of approximately 20% AF^35^. Therefore, we were only able to confirm the diagnosis of IP and the location of the variant on *IKBKG* by applying long-read PCR and Sanger-Sequencing. However, we furthermore employed ONT sequencing and haplotype mapping of methylation profiles in order to investigate ONT’s potential as a possible diagnostic strategy in IP. To our knowledge, this is the first time ONT sequencing has been employed this way, opening up the possibility for its implementation as a second tier (after WES) or even a first tier diagnostic strategy when suspecting IP (and potentially further X-chromosomal dominant disorders associated with skewing of XCI). ONT sequencing can provide two separate strands of information that support the diagnosis without the need for additional analyses, such as traditional XCI-skewing analysis or RNA analyses, thereby saving both time and money in the process. Further analyses reiterating our findings are necessary.

This study underlines the importance of thorough investigation, exceeding the mere variant detection tools of standard genomics softwares, especially in cases with specific phenotypes and underlying genes with highly homologous pseudogenes. It also demonstrates for the first time, the potential of ONT sequencing as a tool for precise variant mapping in the context of differentially methylated promoter regions due to skewed XCI.

## Data Availability

All data produced in the present study are available upon reasonable request to the authors

## Acknowledgements

We thank the family participating in this study, as well as all involved physicians. We also thank the two external laboratories congenics Humangenetik MVZ in Osnabrück, Germany as well as the Institute of Human Genetics, Otto-von-Guericke-Universität Magdeburg, Germany that conducted the long-read sequencing and the X-Inactivation analyses respectively.

## Author’s Contributions

SA, DP and IS conceptualized the project and wrote the manuscript, SA designed the figures, MF and VS were the physicians in charge of the family, HO, MK, DP, BP and IS performed the experiments and interpreted the data. All authors contributed to revision of the manuscript and approved the final version for submission.

## References

(1) Orphanet Report Series. Prevalence and incidence of rare diseases: Bibliographic data. https://www.orpha.net/orphacom/cahiers/docs/GB/Prevalence_of_rare_diseases_by_alphabetical_list.pdf.

(2) Smahi, A.; Courtois, G.; Vabres, P.; Yamaoka, S.; Heuertz, S.; Munnich, A.; Israël, A.; Heiss, N. S.; Klauck, S. M.; Kioschis, P.; Wiemann, S.; Poustka, A.; Esposito, T.; Bardaro, T.; Gianfrancesco, F.; Ciccodicola, A.; D’Urso, M.; Woffendin, H.; Jakins, T.; Donnai, D.; Stewart, H.; Kenwrick, S. J.; Aradhya, S.; Yamagata, T.; Levy, M.; Lewis, R. A.; Nelson, D. L. Genomic rearrangement in NEMO impairs NF-kappaB activation and is a cause of incontinentia pigmenti. The International Incontinentia Pigmenti (IP) Consortium. Nature 2000, 405 (6785), 466–472. DOI: 10.1038/35013114.

(3) Kenwrick, S.; Woffendin, H.; Jakins, T.; Shuttleworth, S. G.; Mayer, E.; Greenhalgh, L.; Whittaker, J.; Rugolotto, S.; Bardaro, T.; Esposito, T.; D’Urso, M.; Soli, F.; Turco, A.; Smahi, A.; Hamel-Teillac, D.; Lyonnet, S.; Bonnefont, J. P.; Munnich, A.; Aradhya, S.; Kashork, C. D.; Shaffer, L. G.; Nelson, D. L.; Levy, M.; Lewis, R. A. Survival of male patients with incontinentia pigmenti carrying a lethal mutation can be explained by somatic mosaicism or Klinefelter syndrome. American journal of human genetics 2001, 69 (6), 1210–1217. DOI: 10.1086/324591. Published Online: Oct. 22, 2001.

(4) Greene-Roethke, C. Incontinentia Pigmenti: A Summary Review of This Rare Ectodermal Dysplasia With Neurologic Manifestations, Including Treatment Protocols. Journal of pediatric health care : official publication of National Association of Pediatric Nurse Associates & Practitioners 2017, 31 (6), e45–e52. DOI: 10.1016/j.pedhc.2017.07.003. Published Online: Sep. 1, 2017.

(5) Parrish, J. E.; Scheuerle, A. E.; Lewis, R. A.; Levy, M. L.; Nelson, D. L. Selection against mutant alleles in blood leukocytes is a consistent feature in Incontinentia Pigmenti type 2. Human molecular genetics 1996, 5 (11), 1777–1783. DOI: 10.1093/hmg/5.11.1777.

(6) Hayden, M. S.; Ghosh, S. Signaling to NF-kappaB. Genes & development 2004, 18 (18), 2195–2224. DOI: 10.1101/gad.1228704.

(7) How, K. N.; Leong, H. J. Y.; Pramono, Z. A. D.; Leong, K. F.; Lai, Z. W.; Yap, W. H. Uncovering incontinentia pigmenti: From DNA sequence to pathophysiology. Frontiers in pediatrics 2022, 10, 900606. DOI: 10.3389/fped.2022.900606. Published Online: Sep. 6, 2022.

(8) Machado, F. B.; Machado, F. B.; Faria, M. A.; Lovatel, V. L.; Da Alves Silva, A. F.; Radic, C. P.; Brasi, C. D. de; Rios, Á. F. L.; Sousa Lopes, S. M. C. de; Da Silveira, L. S.; Ruiz-Miranda, C. R.; Ramos, E. S.; Medina-Acosta, E. 5meCpG epigenetic marks neighboring a primate-conserved core promoter short tandem repeat indicate X-chromosome inactivation. PloS one 2014, 9 (7), e103714. DOI: 10.1371/journal.pone.0103714. Published Online: Jul. 31, 2014.

(9) Allen, R. C.; Zoghbi, H. Y.; Moseley, A. B.; Rosenblatt, H. M.; Belmont, J. W. Methylation of HpaII and HhaI sites near the polymorphic CAG repeat in the human androgen-receptor gene correlates with X chromosome inactivation. American journal of human genetics 1992, 51 (6), 1229–1239.

(10) Fusco, F.; Paciolla, M.; Napolitano, F.; Pescatore, A.; D’Addario, I.; Bal, E.; Lioi, M. B.; Smahi, A.; Miano, M. G.; Ursini, M. V. Genomic architecture at the Incontinentia Pigmenti locus favours de novo pathological alleles through different mechanisms. Human molecular genetics 2012, 21 (6), 1260–1271. DOI: 10.1093/hmg/ddr556. Published Online: Nov. 25, 2011.

(11) Bardaro, T.; Falco, G.; Sparago, A.; Mercadante, V.; Gean Molins, E.; Tarantino, E.; Ursini, M. V.; D’Urso, M. Two cases of misinterpretation of molecular results in incontinentia pigmenti, and a PCR-based method to discriminate NEMO/IKKgamma dene deletion. Human mutation 2003, 21 (1), 8–11. DOI: 10.1002/humu.10150.

(12) Fusco, F.; Pescatore, A.; Conte, M. I.; Mirabelli, P.; Paciolla, M.; Esposito, E.; Lioi, M. B.; Ursini, M. V. EDA-ID and IP, two faces of the same coin: how the same IKBKG/NEMO mutation affecting the NF-κB pathway can cause immunodeficiency and/or inflammation. International reviews of immunology 2015, 34 (6), 445–459. DOI: 10.3109/08830185.2015.1055331. Published Online: Aug. 13, 2015.

(13) Frans, G.; Meert, W.; van der Werff Ten Bosch, J.; Meyts, I.; Bossuyt, X.; Vermeesch, J. R.; Hestand, M. S. Conventional and Single-Molecule Targeted Sequencing Method for Specific Variant Detection in IKBKG while Bypassing the IKBKGP1 Pseudogene. The Journal of molecular diagnostics : JMD 2018, 20 (2), 195–202. DOI: 10.1016/j.jmoldx.2017.10.005. Published Online: Dec. 18, 2017.

(14) Fusco, F.; Pescatore, A.; Steffann, J.; Bonnefont, J.-P.; Oliveira, J. de; Lioi, M. B.; Ursini, M. V. Clinical utility gene card: for incontinentia pigmenti. European journal of human genetics : EJHG 2019, 27 (12), 1894–1900. DOI: 10.1038/s41431-019-0463-9. Published Online: Jul. 9, 2019.

(15) Richards, S.; Aziz, N.; Bale, S.; Bick, D.; Das, S.; Gastier-Foster, J.; Grody, W. W.; Hegde, M.; Lyon, E.; Spector, E.; Voelkerding, K.; Rehm, H. L. Standards and guidelines for the interpretation of sequence variants: a joint consensus recommendation of the American College of Medical Genetics and Genomics and the Association for Molecular Pathology. Genetics in medicine : official journal of the American College of Medical Genetics 2015, 17 (5), 405–424. DOI: 10.1038/gim.2015.30. Published Online: Mar. 5, 2015.

(16) Ellard, S.; Baple, E. L.; Berry, I.; Forrester, N.; Turnbull, C.; Owens, M.; Eccles, D. M.; Abbs, S.; Scott, R.; Deans, Z. C.; Lester, T.; Campbell, J.; Newman, W. G.; McMullan, D. J. ACGS Best Practice Guidelines for Variant Classification 2019 2019.

(17) ClinGen Sequence Variant Interpretation Recommendation for PM2 - Version 1.0. https://clinicalgenome.org/site/assets/files/5182/pm2_-_svi_recommendation_-_approved_sept2020.pdf.

(18) Landrum, M. J.; Lee, J. M.; Benson, M.; Brown, G. R.; Chao, C.; Chitipiralla, S.; Gu, B.; Hart, J.; Hoffman, D.; Jang, W.; Karapetyan, K.; Katz, K.; Liu, C.; Maddipatla, Z.; Malheiro, A.; McDaniel, K.; Ovetsky, M.; Riley, G.; Zhou, G.; Holmes, J. B.; Kattman, B. L.; Maglott, D. R. ClinVar: improving access to variant interpretations and supporting evidence. Nucleic acids research 2018, 46 (D1), D1062–D1067. DOI: 10.1093/nar/gkx1153.

(19) Bolduc, V.; Chagnon, P.; Provost, S.; Dubé, M.-P.; Belisle, C.; Gingras, M.; Mollica, L.; Busque, L. No evidence that skewing of X chromosome inactivation patterns is transmitted to offspring in humans. The Journal of clinical investigation 2008, 118 (1), 333–341. DOI: 10.1172/JCI33166.

(20) Zheng, Z.; Li, S.; Su, J.; Leung, A. W.-S.; Lam, T.-W.; Luo, R. Symphonizing pileup and full-alignment for deep learning-based long-read variant calling. Nat Comput Sci 2022, 2 (12), 797–803. DOI: 10.1038/s43588-022-00387-x.

(21) Cheetham, S. W.; Kindlova, M.; Ewing, A. D. Methylartist: tools for visualizing modified bases from nanopore sequence data. Bioinformatics (Oxford, England) 2022, 38 (11), 3109–3112. DOI: 10.1093/bioinformatics/btac292.

(22) Voigt M, Fusch C, Olbertz D et al. Analyse des Neugeborenenkollektivs der Bundesrepublik Deutschland. 12. Mitteilung: Vorstellung engmaschiger Perzentilwerte (-kurven) für die Körpermaße Neugeborener. In Geburtsh Frauenheilk 66; pp 956–970.

(23) Kromeyer-Hauschild, K., Wabitsch, M., Kunze, D. et al. Perzentile für den Body-mass-Index für das Kindesund Jugendalter unter Heranziehung verschiedener deutscher Stichproben. In Monatsschr Kinderheilkd 149; pp 807–818.

(24) Berlin, A. L.; Paller, A. S.; Chan, L. S. Incontinentia pigmenti: a review and update on the molecular basis of pathophysiology. Journal of the American Academy of Dermatology 2002, 47 (2), 169–87; quiz 188-90. DOI: 10.1067/mjd.2002.125949.

(25) Poziomczyk, C. S.; Recuero, J. K.; Bringhenti, L.; Maria, F. D. S.; Campos, C. W.; Travi, G. M.; Freitas, A. M.; Maahs, M. A. P.; Zen, P. R. G.; Fiegenbaum, M.; Almeida, S. T. de; Bonamigo, R. R.; Bau, A. E. K. Incontinentia pigmenti. Anais brasileiros de dermatologia 2014, 89 (1), 26–36. DOI: 10.1590/abd1806-4841.20142584.

(26) Fusco, F.; Bardaro, T.; Fimiani, G.; Mercadante, V.; Miano, M. G.; Falco, G.; Israël, A.; Courtois, G.; D’Urso, M.; Ursini, M. V. Molecular analysis of the genetic defect in a large cohort of IP patients and identification of novel NEMO mutations interfering with NF-kappaB activation. Human molecular genetics 2004, 13 (16), 1763–1773. DOI: 10.1093/hmg/ddh192. Published Online: Jun. 30, 2004.

(27) Dangouloff-Ros, V.; Hadj-Rabia, S.; Oliveira Santos, J.; Bal, E.; Desguerre, I.; Kossorotoff, M.; An, I.; Smahi, A.; Bodemer, C.; Munnich, A.; Steffann, J.; Boddaert, N. Severe neuroimaging anomalies are usually associated with random X inactivation in leucocytes circulating DNA in X-linked dominant Incontinentia Pigmenti. Molecular genetics and metabolism 2017, 122 (3), 140–144. DOI: 10.1016/j.ymgme.2017.07.001. Published Online: Jul. 10, 2017.

(28) Sharp, A.; Robinson, D.; Jacobs, P. Age- and tissue-specific variation of X chromosome inactivation ratios in normal women. Human genetics 2000, 107 (4), 343–349. DOI: 10.1007/s004390000382.

(29) de Jesus A, Torreggiani S, Lin B, Mitchell J, Karlins E, Oler A, Alehashemi S, Kahle D, Honer K, Souto Adeva G, Hanson E, Montealegre Sanchez G, Khojah A, Moran T, Wu E, Scott C, Leahy T, MacDermott E, Killeen O, Arkachaisri T, Gucev Z, Phillippi K, Mammadova V, Nasrullayeva G, Goldbach-Mansky R., Ed. Splice Site Variants in IKBKG, Encoding NEMO, Detected by a Customized Analysis of Next-Generation Sequencing Data Cause an Earlyonset Autoinflammatory Syndrome of Panniculitis and Cytopenias in Male and Female Patients; Arthritis Rheumatol., 2020.

(30) Johansson, J.; Lidéus, S.; Höijer, I.; Ameur, A.; Gudmundsson, S.; Annerén, G.; Bondeson, M.-L.; Wilbe, M. A novel quantitative targeted analysis of X-chromosome inactivation (XCI) using nanopore sequencing. Scientific reports 2023, 13 (1), 12856. DOI: 10.1038/s41598-023-34413-3. Published Online: Aug. 8, 2023.

(31) Losson, R.; Lacroute, F. Interference of nonsense mutations with eukaryotic messenger RNA stability. Proceedings of the National Academy of Sciences of the United States of America 1979, 76 (10), 5134–5137. DOI: 10.1073/pnas.76.10.5134.

(32) Kurosaki, T.; Popp, M. W.; Maquat, L. E. Quality and quantity control of gene expression by nonsense-mediated mRNA decay. Nature reviews. Molecular cell biology 2019, 20 (7), 406–420. DOI: 10.1038/s41580-019-0126-2.

(33) Noensie, E. N.; Dietz, H. C. A strategy for disease gene identification through nonsense-mediated mRNA decay inhibition. Nature biotechnology 2001, 19 (5), 434–439. DOI: 10.1038/88099.

(34) Häuser, F.; Gökce, S.; Werner, G.; Danckwardt, S.; Sollfrank, S.; Neukirch, C.; Beyer, V.; Hennermann, J. B.; Lackner, K. J.; Mengel, E.; Rossmann, H. A non-invasive diagnostic assay for rapid detection and characterization of aberrant mRNA-splicing by nonsense mediated decay inhibition. Molecular genetics and metabolism 2020, 130 (1), 27–35. DOI: 10.1016/j.ymgme.2020.03.002. Published Online: Mar. 19, 2020.

(35) Lin, M.-T.; Mosier, S. L.; Thiess, M.; Beierl, K. F.; Debeljak, M.; Tseng, L.-H.; Chen, G.; Yegnasubramanian, S.; Ho, H.; Cope, L.; Wheelan, S. J.; Gocke, C. D.; Eshleman, J. R. Clinical validation of KRAS, BRAF, and EGFR mutation detection using next-generation sequencing. American journal of clinical pathology 2014, 141 (6), 856–866. DOI: 10.1309/AJCPMWGWGO34EGOD.

(36) Pescatore, A.; Spinosa, E.; Casale, C.; Lioi, M. B.; Ursini, M. V.; Fusco, F. Human Genetic Diseases Linked to the Absence of NEMO: An Obligatory Somatic Mosaic Disorder in Male. International journal of molecular sciences 2022, 23 (3). DOI: 10.3390/ijms23031179. Published Online: Jan. 21, 2022.

