## Supplementary Material for "Incorporating Nanopore Sequencing into a Diverse Diagnostic Toolkit for Incontinentia Pigmenti"

**Table 1:** Primer-pairs used for bidirectional Sanger-Sequencing

| Primer | Sequence | Genomic position (GRCh38) on ChrX | Length of PCR product in bp |
|---|---|---|---|
| Exon 3.1 Forward Primer | 5’-GAGCTTCTGCATTTCCAAGC-3’ | 154,556,209 | 324 |
| Exon 3.1 Reverse Primer | 5’-TAAACCGGAACTGGGAGTGT-3’ | 145,556,532 | |
| Exon 3.2 Forward Primer | 5’-CTGGCCTCTGACTTCCTGAG-3’ | 154,556,066 | 392 |
| Exon 3.2 Reverse Primer | 5’-TTGGGGGACCCCGACCACGG-3’ | 154,556,457 | |

**Table 2:** RNA-Primer used for bidirectional Sanger-Sequencing

| Primer | Sequence | cDNA position on NM_001099857.5 | Exon | Length of PCR product in bp |
|---|---|---|---|---|
| RNA Primer B F | 5’-CGGCAGCAGATCAGGACGTA-3’ | c.59 | 2 | 347 |
| RNA Primer B R | 5’-CATCTGCTGCTGGCATCTCT-3’ | c.405 | 3/4 | |
| RNA Primer C F | 5’-GGCACCTCTGGAAGAGCCAAC-3’ | c.8 | 2 | 390 |
| RNA Primer C R | 5’-GCTGGCATCTCTTCAGGTGCT-3’ | c.397 | 3 | |

**Figure S1:** (Created with BioRender.com) Pedigree; affected female individual, mother and maternal grandmother with “skin lesions”; mother and index are both carriers of the *IKBKG* variant, the maternal grandmother was not tested

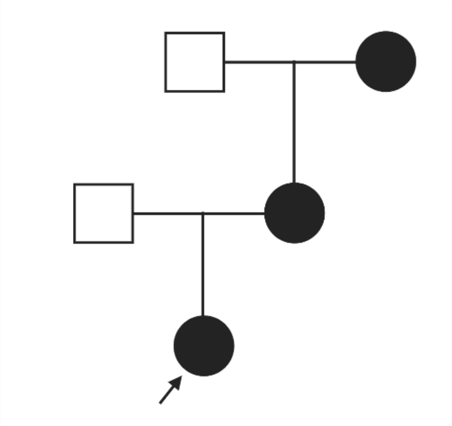

**Figure S2:** from Integrative Genomics Viewer; Viewing of the *IKBKG* locus and the detected 5 bp deletion in IGV. Transparent reads specify mapping quality of 0 indicating that reads can not be mapped conclusively to a single region.

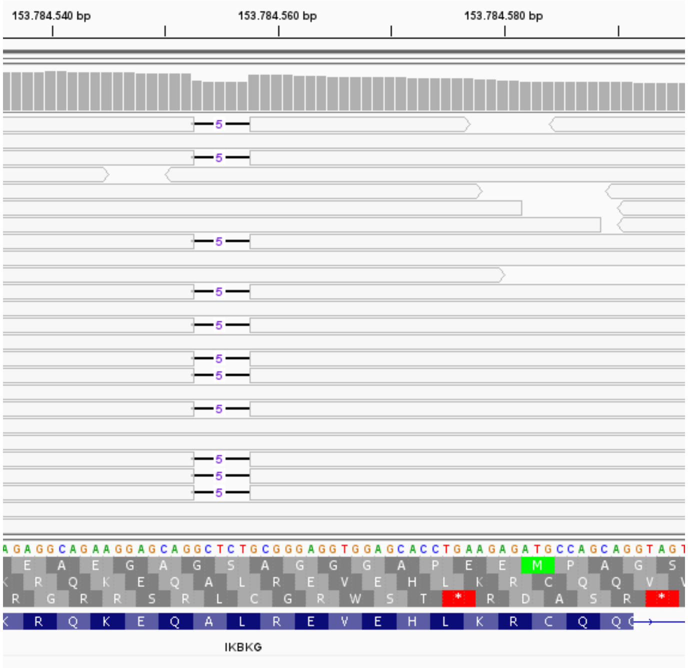

### Methylation status of the *RP2* and *AR* loci

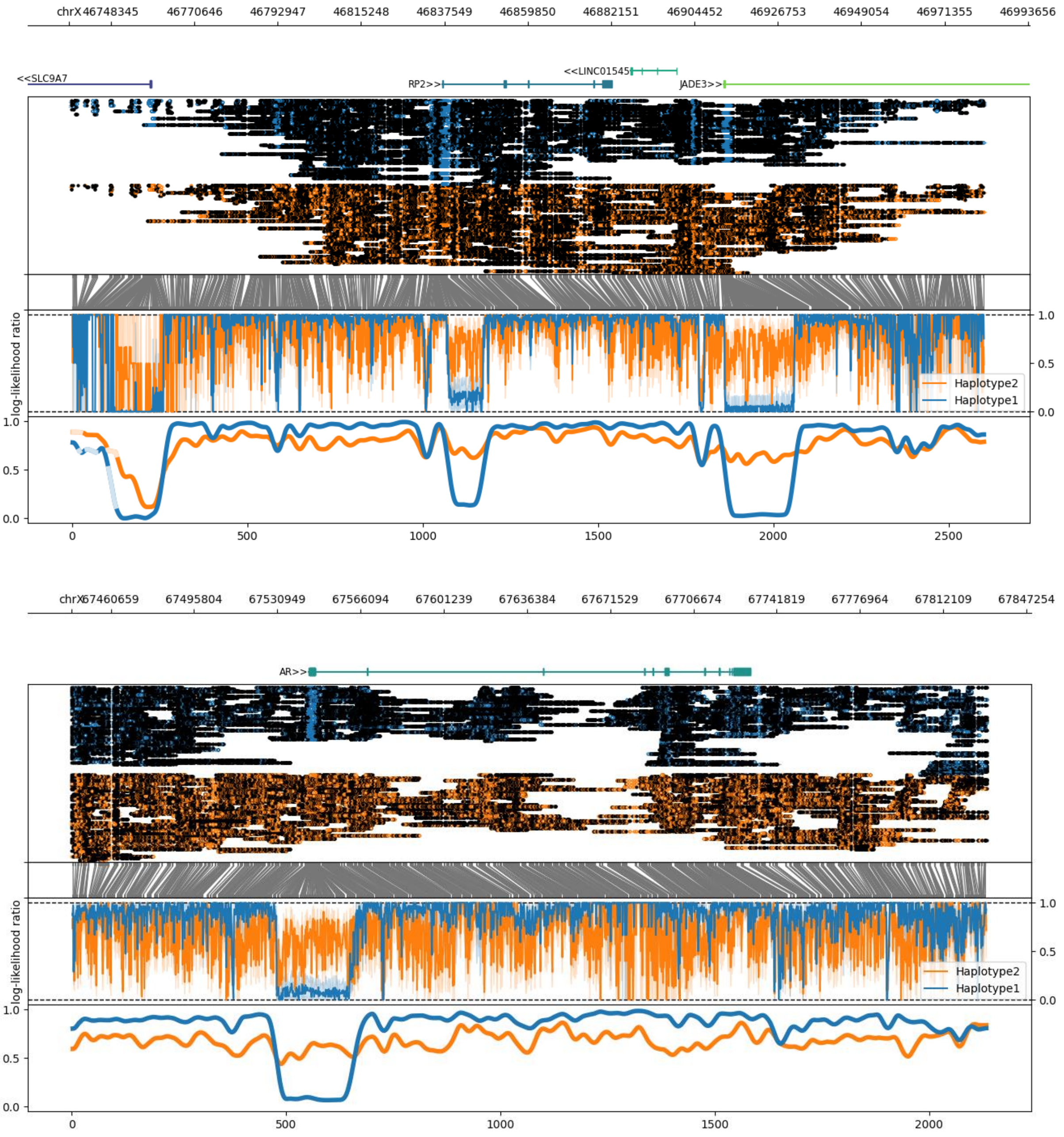


**Figure S3:** Haplotype-specific methylation profiles of the *RP2* (a) and the *AR* (b) locus showing differentially methylated haplotypes. In case of random XCI a similar methylation of both haplotypes would be expected. From top to bottom these plots show a gene track, haplotype-specific methylation calls relative to aligned read positions, a translation from genome space into a modified base space consisting only of instances of the methylated motif, the haplotype-specific methylation statistic (log-likelihood ratio) and a smoothed sliding-window plot showing methylation fraction across the region (*RP2/AR* gene ± 100 kb).
